# Conventional electrode catheter placement can miss crucial atypical atrioventricular nodal reentrant tachycardia circuit details. -New insights into the retrograde slow pathways-

**DOI:** 10.1101/2024.01.16.24301387

**Authors:** Mihoko Kawabata, Shingo Maeda, Kaoru Okishige, Yasuhiro Shirai, Tatsuaki Kamata, Tomoyuki Kawashima, Ryo Yonai, Hirotsugu Atarashi, Kenzo Hirao

**Affiliations:** Heart Rhythm Center, AOI Universal Hospital, Kanagawa, Japan; Yokohama Minato Heart Clinic, Kanagawa, Japan; Heart Rhythm Center, National Hospital Organization Disaster Medical Center, Tokyo, Japan; Division of Cardiovascular Disease, AOI Universal Hospital, Kanagawa, Japan

**Keywords:** atypical atrioventricular nodal reentrant tachycardia, atrioventricular node inferior extensions, tricuspid annulus, coronary sinus, electroanatomical mapping

## Abstract

**Background:** During atypical atrioventricular nodal reentrant tachycardia (AVNRT), the earliest atrial activation site following retrograde slow pathway (SP) conduction is at the atrial exit of the left inferior extension of the compact node (LIE) in the coronary sinus (CS) or the right inferior extension (RIE) on the tricuspid annulus (TA). We tested the validity of conventional electrode placement-based mapping of the atrial ends of these extensions.

**Methods:** We retrospectively evaluated the efficiency of the two catheter (His bundle and CS) mapping method for localization of LIE and RIE in atypical AVNRT patient using electroanatomical 3D mapping validation.

**Results:** Among 19 atypical AVNRTs (15 fast/slow and 4 slow/slow) in 14 patients (9 females, age 59±17), 8 AVNRTs had LIE involvement and 11 had RIE. The 8 LIE exits were inside the CS, and localization by 3D mapping and CS electrode catheter matched in all. In contrast, RIE exits were on the posterior TA where electrode catheters are conventionally not placed. All RIE exits required 3D mapping for accurate localization. During retrograde RIE conduction, comparison of the activation time of the CS ostium and HBE showed that the CS ostium was earlier in 7 RIEs, HBE was earlier in 1, and they were simultaneous in 3, resulting in the presence of RIE being missed in 4/11 (36%) AVNRTs using current diagnostic criteria. Activation time of the CS ostium and His bundle were determined by their relative closeness to the RIE exit.

**Conclusions:** Conventionally placed electrode catheter mapping in atypical AVNRT was able to identify 100% of LIE, but only 64% of RIE. It is critical to place a catheter on or use a 3D mapping system for the posterior TA in cases of suspected atypical AVNRT, so that all inferior extensions of the AV node can be identified and targeted for treatment.

## Introduction

Catheter ablation is the current treatment of choice in patients with symptomatic atrioventricular nodal reentrant tachycardia (AVNRT). Successful cure of AVNRT is now possible in nearly all patients. However, the reentrant circuit of AVNRT is still not completely understood^1–5^. In cases of atypical AVNRT, retrograde conduction in the slow pathway (SP) is classically diagnosed when ventriculoatrial (VA) conduction time jumps (increases abruptly) during electrophysiological (EP) study, associated with a change in retrograde atrial activation sequence, namely, the earliest retrograde atrial activation site shifts from the His bundle area to the ostium of the coronary sinus (CS)^6^.

Histologically, AV nodal SPs are believed to correspond to the right and left inferior extensions (RIE and LIE) of the compact AV node that connect to atrial cardiomyocytes at the atrial end of the SP^7,8^. Classically, the ablation target for atypical AVNRT is the same SP as for typical AVNRT, in the posterior right atrial septum.

We have previously reported based on electroanatomical 3D mapping, that the sites of the earliest retrograde atrial activation during atypical AVNRT were either inside the CS, corresponding to the location of the exit of the LIE, or in the posterior tricuspid annulus (TA), corresponding to the location of the exit of the RIE^1^. Thus, LIE is easily detected by the CS catheter, but RIE would usually be overlooked due to absence of catheters at the posterior TA. The purpose of the current study was to test the validity of conventional electrode placement-based mapping of the atrial end of LIE and RIE functioning as retrograde SP during atypical AVNRT.

## Methods

### Patients

All patients who underwent an EP study and catheter ablation for atypical AVNRT between April 2021 and June 2023 at our institution were retrospectively identified and included in the study. The study received approval from our institutional review board, and all patients provided written, informed consent.

### EP study

All patients were studied in the postabsorptive state under mild sedation and after all antiarrhythmic agents had been discontinued for at least four half-lives. No patient had received amiodarone in the preceding 3 months. Two quadripolar catheters were introduced from the femoral veins and positioned at the high right atrium (HRA) and right ventricular apex (RVA). A deflectable decapolar electrode catheter with 2-mm interelectrode spacing (Abbott Laboratories, Chicago, Illinois, USA) was positioned within the CS and a hexapolar electrode catheter with 2-/7-/5-/5-/5-mm inter-electrode spacing (Japan Lifeline Co., Japan) in the His bundle region (HBE) (Figure 1). This electrode catheter placement is conventional for this type of EP study for AVNRT. The proximal bipole of the CS catheter was located near the CS ostium, which was confirmed by retrograde CS venography.

**Figure 1.**
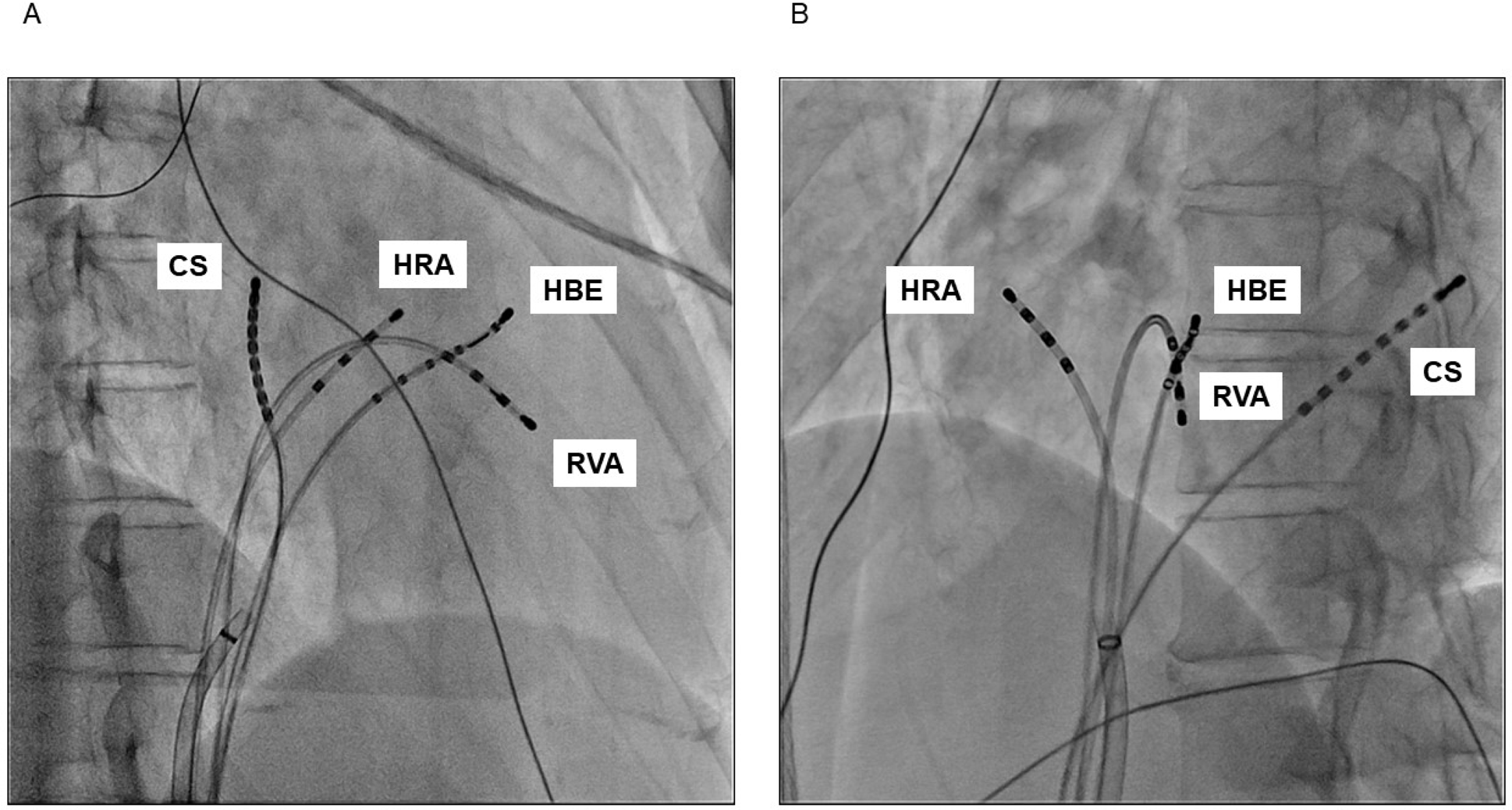
Fluoroscopic images of the conventional electrode catheter setting for EP study for AVNRT in the right anterior oblique (RAO; panel A) and left anterior oblique (LAO; panel B) projections. Catheters are positioned at the coronary sinus (CS), high right atrium (HRA), right ventricular apex (RVA), and His bundle electrogram region (HBE). The CS catheter was withdrawn such that the proximal pair was at the ostium of the CS.

Baseline EP study included atrial or ventricular programmed stimulation (burst or extrastimulus pacing). If tachycardia was not induced, the same pacing maneuvers were repeated under isoproterenol infusion (0.5-2.0 µg/min). AVNRT was diagnosed according to published criteria and pacing maneuvers as applicable^6,9^. Standard definitions of the mechanism of AVNRT were used^2,6^. The presence of an accessory pathway was excluded when the VA interval during the tachycardia was not lengthened by an occurrence of bundle branch block; para-Hisian pacing during sinus rhythm exhibited a retrograde AV nodal conduction pattern^10^; and a positive response to the delivery of His-refractory ventricular premature depolarization during tachycardia was absent. Positive responses include: (1) termination with VA block, (2) resetting with advancement, and (3) resetting with delay. Atrial tachycardia was excluded when a V-A-V sequence (not a V-A-A-V sequence) was observed on cessation of ventricular pacing associated with 1:1 VA conduction during the tachycardia and the tachycardia was reproducibly terminated with ventricular extrastimuli not reaching the atrium^11^. Retrograde fast pathway (FP) conduction was assumed when the earliest retrograde atrial activation site was on the HBE catheter trace during ventricular pacing^6^. Retrograde SP conduction was defined when the earliest retrograde atrial activation site was not the HBE catheter or when the retrograde atrial activation sequence in which the HBE catheter was the earliest site shifted to other sites^6,11^. VA conduction jump was defined using the criterion of an increase of at least 50 ms in the VA interval for a 10 ms decrease in the ventricular extrastimulus coupling interval^2,6^.

After the conventionally placed electrode catheter mapping was performed, the retrograde atrial activation sequence during atypical AVNRT or ventricular pacing was determined by electroanatomical 3D mapping (CARTO®, Biosense Webster, Diamond Bar, California, USA or Ensite™, Abbott Laboratories, Chicago, Illinois, USA) using PentaRay (Biosense Webster) or HD Grid mapping catheters (Abbott Laboratories) as previously described^1^ with particular attention paid to the CS ostium, tricuspid valve, and His bundle areas. The extent of the region where the His potential was recordable was demarcated. The distance between 2 points was measured on the 3D map. When measuring distances from the CS ostium, the most inferior point (floor) of the CS ostium rim was used as the reference point.

For the purposes of this paper, we denote a retrograde SP with LIE exit as ‘slow (LIE)’, and a retrograde SP with RIE exit as ‘slow (RIE)’.

### Catheter ablation

Applications of radiofrequency energy were made using a 4-mm tip ablation catheter (THERMOCOOL SMARTTOUCH catheter, Biosense Webster, or FlexAbility ablation catheter, Abbott Laboratories). Our targets were either the earliest retrograde atrial activation site during tachycardia or ventricular pacing, or conventional SP targets (posterior right atrial septum) identified by a combined anatomic and electrogram recording approach. For patients who displayed both typical and atypical AVNRT, the target of ablation was the conventional SP region. Electrograms and ablation characteristics were examined, and the anatomic location of the successful ablation site was noted on the 3D map. Procedural endpoints were reached when aggressive atrial and ventricular programmed stimulation before and after administration of intravenous isoproterenol produced no more than a single AV nodal echo.

Data are expressed as the mean ± standard deviation.

## Results

Fourteen patients (9 women; age, 59±17 years; left ventricular ejection fraction, 60±12 %) with a total of 19 atypical AVNRTs (15 fast/slow and 4 slow/slow types) who underwent EP study and ablation during the study period were included in this study. No patient had structural heart disease. Two patients also had slow/fast tachycardia, i.e., typical AVNRT.

### VA conduction through SP in the patients with atypical AVNRT

In 2 patients, AVNRT occurred spontaneously or was induced easily, therefore, EP study could not be performed thoroughly in them. Six patients showed classical retrograde dual VA conduction physiology, in whom a VA jump was detected in 3, during the shift from FP to SP. Among these 6 patients, the earliest retrograde atrial activation site shifted from HBE to the CS ostium in 2, to inside the CS in 2, and to simultaneous HBE and CS ostium activation in 2. Four patients had no retrograde FP conduction but only SP conduction at baseline.

In the remaining 2 patients, there was no VA conduction under ventricular pacing, but AVNRT frequently began spontaneously, or was induced by atrial pacing. In both patients the tachycardia was reproducibly terminated with ventricular extrastimuli not reaching the atrium.

In the 12 patients in whom VA conduction was present and could be studied, the retrograde atrial activation sequence during the tachycardia was identical to that during ventricular pacing through the retrograde SP. Para-Hisian pacing during sinus rhythm was performed in 9, exhibiting a retrograde AV nodal conduction pattern in all. The tachycardia was not reset by ventricular extrastimuli delivered while the His bundle was refractory in all 10 in whom that was examined. In the end, among 14 patients, there was a sole LIE exit site in 4, a sole RIE exit site in 6, and dual LIE and RIE exit sites in 4 patients.

### The retrograde atrial exit sites of atypical AVNRT

The 4 patients with dual SP exit sites consisted of 2 patients who displayed both fast/slow (LIE) and fast/slow (RIE) and 2 in whom fast/slow (RIE) AVNRT became manifest only after the LIE exit of fast/slow (LIE) was ablated. One patient exhibited fast/slow (RIE) and slow/slow (RIE) AVNRT. As a result, among 19 atypical AVNRTs, 8 AVNRTs had LIE involvement and 11 had RIE involvement. (Table 1)

**Table 1.**
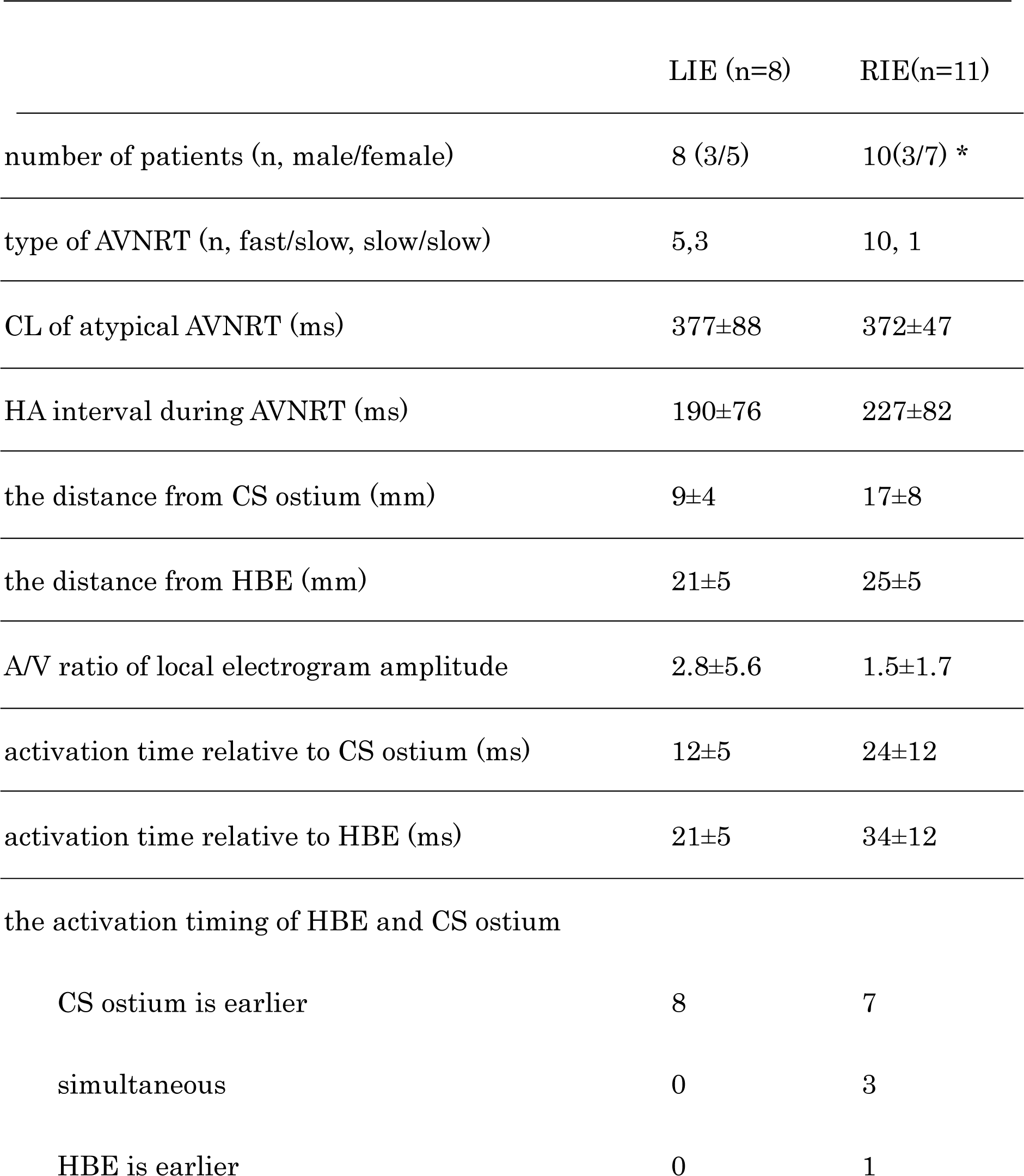
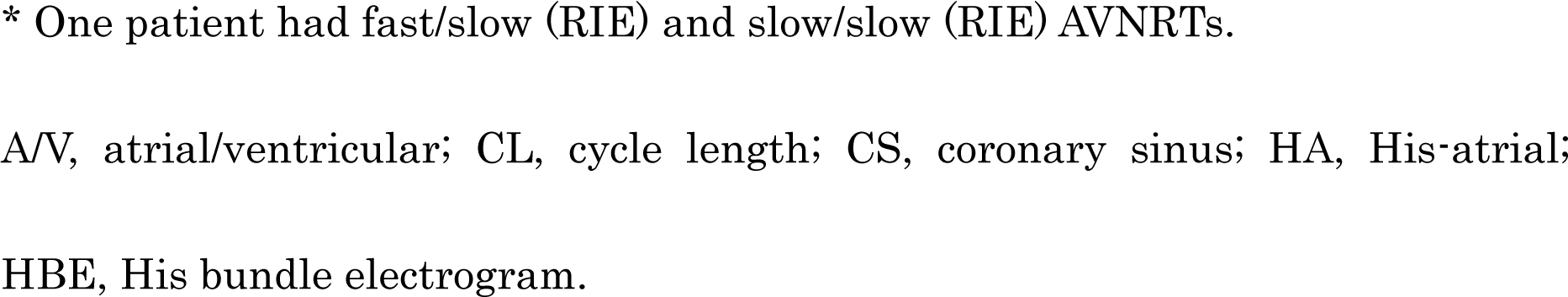
Clinical characteristics of the exits of left inferior extension (LIE) and right inferior extension (RIE) during 19 atypical AVNRT (atrioventricular nodal reentrant tachycardia)

|  | LIE (n=8) | RIE(n=11) |
| --- | --- | --- |
| number of patients (n, male/female) | 8 (3/5) | 10(3/7) * |
| type of AVNRT (n, fast/slow, slow/slow) | 5,3 | 10, 1 |
| CL of atypical AVNRT (ms) | 377±88 | 372±47 |
| HA interval during AVNRT (ms) | 190±76 | 227±82 |
| the distance from CS ostium (mm) | 9±4 | 17±8 |
| the distance from HBE (mm) | 21±5 | 25±5 |
| A/V ratio of local electrogram amplitude | 2.8±5.6 | 1.5±1.7 |
| activation time relative to CS ostium (ms) | 12±5 | 24±12 |
| activation time relative to HBE (ms) | 21±5 | 34±12 |
| the activation timing of HBE and CS ostium |  |  |
| CS ostium is earlier | 8 | 7 |
| simultaneous | 0 | 3 |
| HBE is earlier | 0 | 1 |

\* One patient had fast/slow (RIE) and slow/slow (RIE) AVNRTs.
A/V, atrial/ventricular; CL, cycle length; CS, coronary sinus; HA, His-atrial;
HBE, His bundle electrogram.

### VA conduction through SP with LIE exit (Table 1, Figure 2)

In all 8 AVNRTs of SP conduction through the LIE (fast/slow (LIE) or slow/slow (LIE)), the earliest retrograde atrial activation sites identified by 3D mapping perfectly matched the sites identified by the CS electrode catheter, and were inside the CS (Figure 2). The atrial conduction sequence recorded by the CS catheter electrodes displayed a focal pattern, resulting in the elucidation of the presence of LIE in 8/8 AVNRTs using the conventional deployment of electrode catheters.

**Figure 2.**
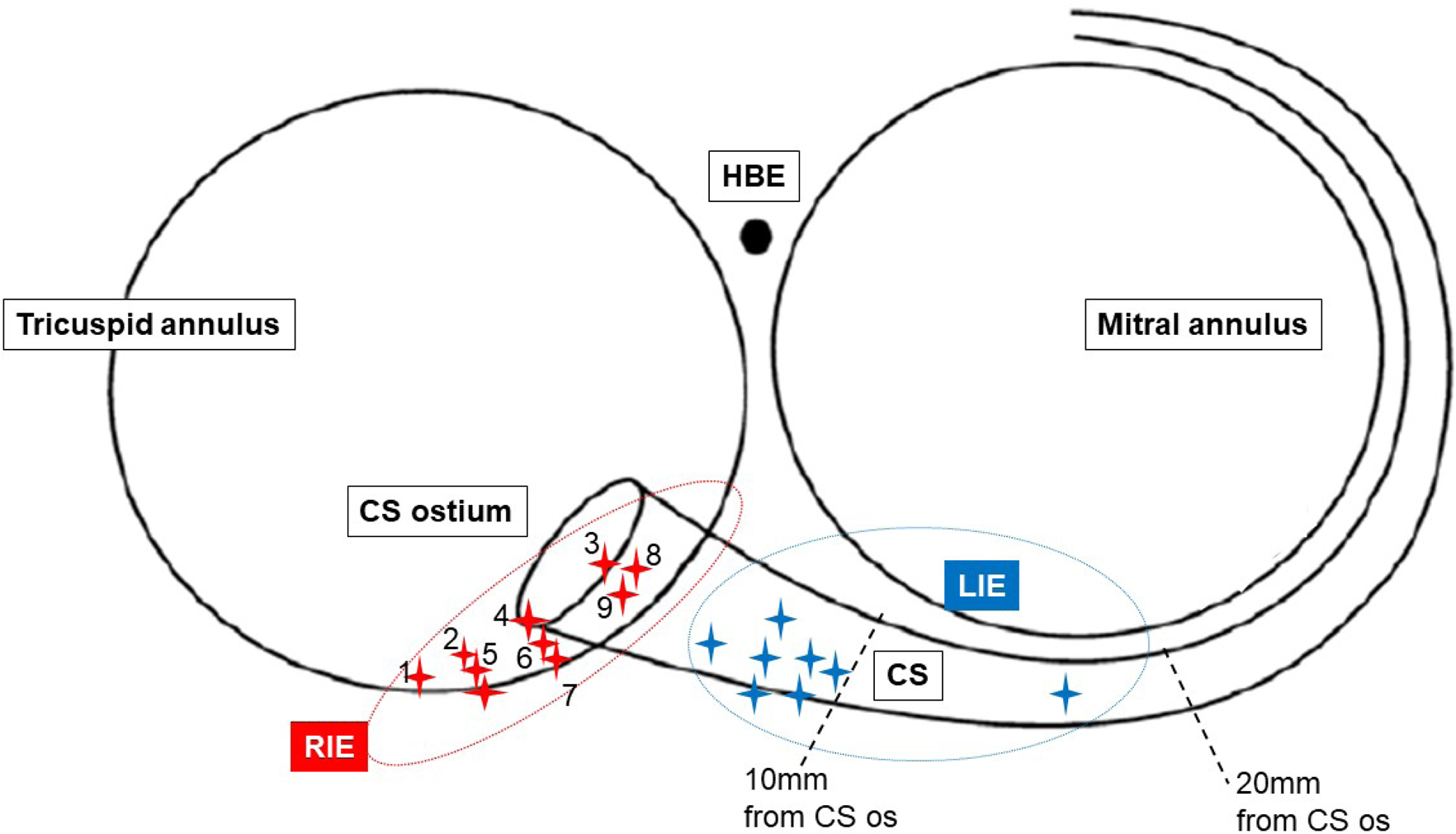
Schematic representation of the location of the earliest atrial activation site during tachycardia or ventricular pacing in atypical AVNRTs, as seen from the left anterior oblique projection. Both fast/slow (LIE) and fast/slow (RIE) AVNRTs are observed in 4 patients. In one patient who had the RIE exit at 5:30 direction without a small number, the location of HBE was missed. In another patient retrograde conduction during slow/slow (RIE) AVNRT was fragile and unmappable. Therefore, the schema represents the location in 18 AVNRTs. All exits of LIE are inside the CS, indicated in blue. The distance from the atrial exits of LIE to the CS ostium was 9±4 mm (4-18mm). RIE exits (indicated in red) were located on the TA posterior to the CS ostium (between 4 and 6 o’clock). The small numbers by the RIE exit site symbol correspond to patient numbers in Figure 5. LIE = leftward inferior extension; RIE = rightward inferior extension. Other abbreviations are as in Figure 1.

VA conduction and anatomical assessment for the SP with LIE exit was performed. The distance from the atrial exits of LIE to the CS ostium was 9±4 mm (range 4-18) and to HBE was 21±5 mm (13-26). The earliest electrogram of LIE preceded that of CS ostium and HBE (by 12±5 msec for the CS ostium and 21±5 msec for the HBE) during the tachycardia or ventricular pacing. Comparison of the local electrogram of the CS ostium and HBE showed that the CS ostium activation was earlier in all 8 cases. Figure 3 shows an example of fast/slow (LIE) AVNRT.

**Figure 3.**
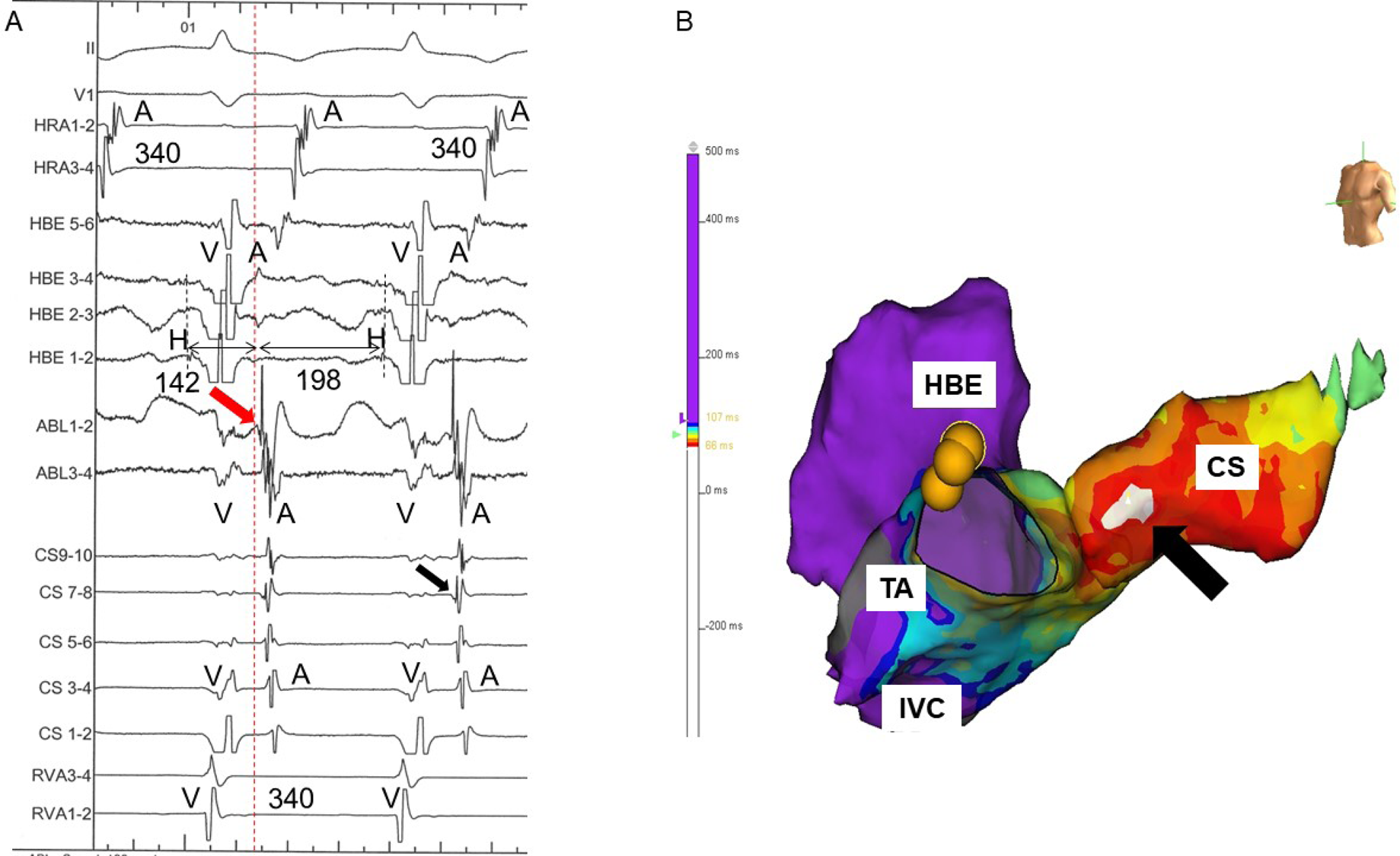
A patient with fast/slow (LIE) AVNRT. Panel A. The electrocardiograms during atypical AVNRT using the LIE for the SP display earliest retrograde atrial activation on electrodes ABL1-2 (red arrow), inside the CS. Among the conventionally positioned catheters, CS7-8 displays the earliest activation (black arrow). Panel B. EnSite mapping of the retrograde atrial activation during AVNRT in the LAO view shows the earliest site (arrow), which is 8 mm distal from CS ostium. Yellow tags indicate the sites where the His bundle potential is recorded. The A/V ratio of local electrogram amplitude of the atrial exit of LIE was 1.6. IVC = inferior vena cava; S1 = stimulus. Other abbreviations are as in the previous figures.

### VA conduction through SP with RIE exit (Table 1, Figure 2)

With respect to the atrial conduction sequence using the conventional deployment of electrode catheters, the atrial end of RIE could not be identified anatomically in all 11 AVNRTs. 3D electroanatomical mapping was necessary to reveal RIE exits located on the TA posterior to the CS ostium (between 4 and 6 o’clock in the left anterior oblique projection). The distance from the atrial exits of RIE to the CS ostium was 17±8 mm (4-28mm) and to HBE was 25±5 mm (19-36mm).

The atrial conduction sequence showed that the earliest electrogram of RIE preceded that of CS ostium and HBE by 24±12 and 34±12 msec respectively. Comparison of the local electrogram of the CS ostium and HBE during atypical AVNRT or ventricular pacing showed that the CS ostium activation was earlier in 7 RIEs, HBE was earlier in 1, and they were simultaneous in 3. Therefore, by current diagnostic criteria, the presence of RIE was not identified in 4 out of 11 (36%) AVNRTs using conventional catheter mapping. Figure 4 shows an example of fast/slow (RIE) AVNRT, in which the atrial electrogram at HBE is ahead of that in CS ostium and the atrial sequence would be regarded as retrograde conduction over FP by classical criteria.

**Figure 4.**
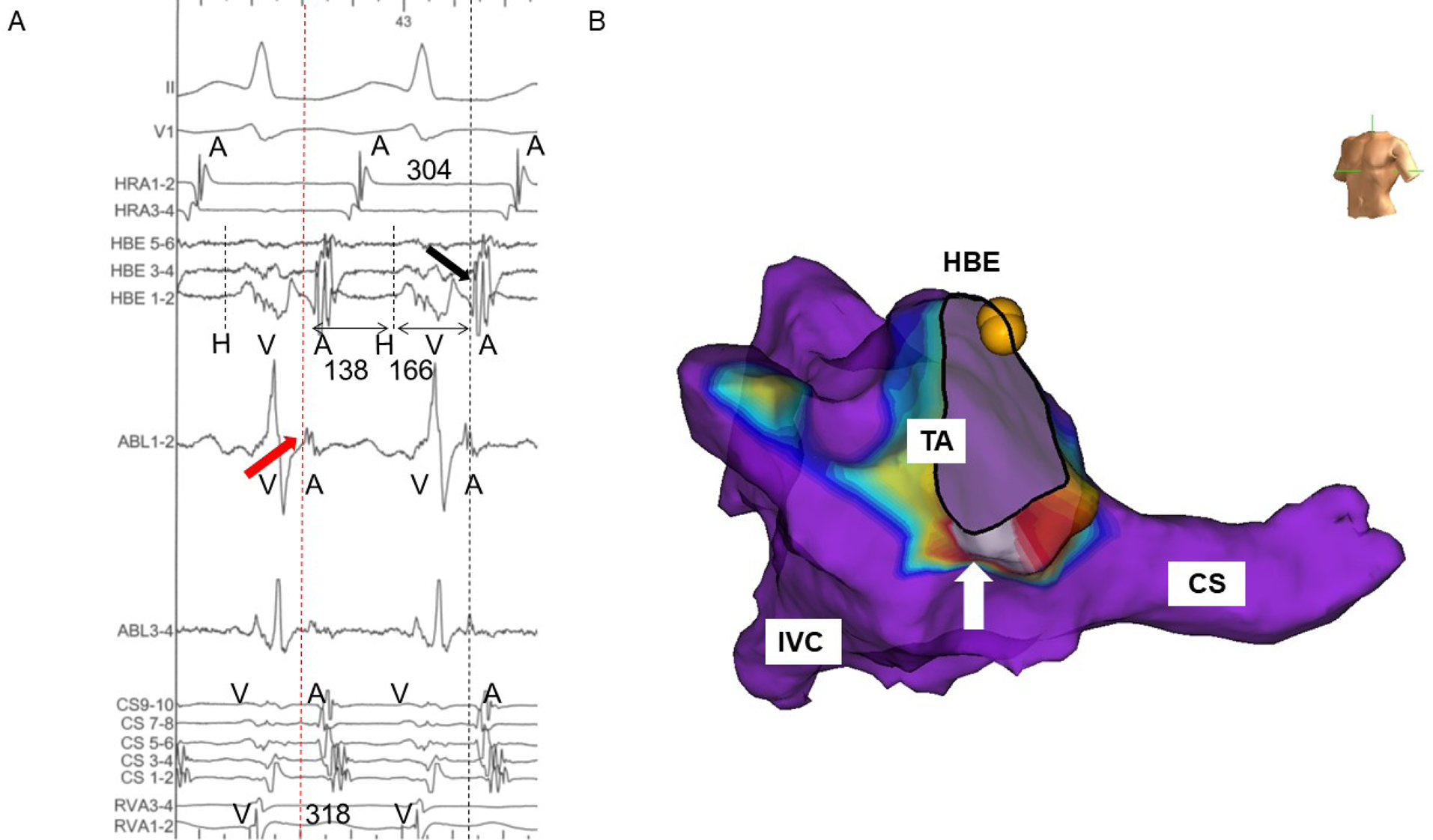
Patient 1: fast/slow (RIE) AVNRT in which the electrogram in HBE was ahead of that in CS ostium. Panel A. The electrocardiograms show that with respect to the atrial conduction sequence using the conventional deployment of electrode catheters, HBE1-2 is 5ms ahead of CS7-8 (black arrow), suggesting that the retrograde pathway might be FP. Therefore, we mapped the aortic cusp to find an earlier activation site that might be a potential target location for safe catheter ablation, but aortic cusp activation was late. Panel B. EnSite mapping of the retrograde atrial activation during AVNRT revealed that the earliest site was on the TA at 6 o’clock projection in the LAO view (arrow), which is the exit site of RIE. The catheter placed on that site (ABL1-2, red arrow) is the earliest in panel A. Yellow tags indicate the sites where the His bundle potential was recorded. HBE is closer to the atrial exit site of RIE with the distance of 19mm compared to the CS ostium, the distance from which to the atrial exit site of RIE was 28mm. The A/V ratio of local electrogram amplitude of the atrial exit site of RIE was 0.23. Abbreviations are as in Figure 1.

We analyzed the temporal and anatomical relationship between HB site and CS ostium. A plot of difference in activation time of HBE and CS ostium vs the difference in their distance from the atrial exit of RIE showed a roughly linear relationship (Figure 5). This graph strongly suggests that proximity to the atrial end of RIE determines how early one or the other is excited relative to the other.

**Figure 5.**
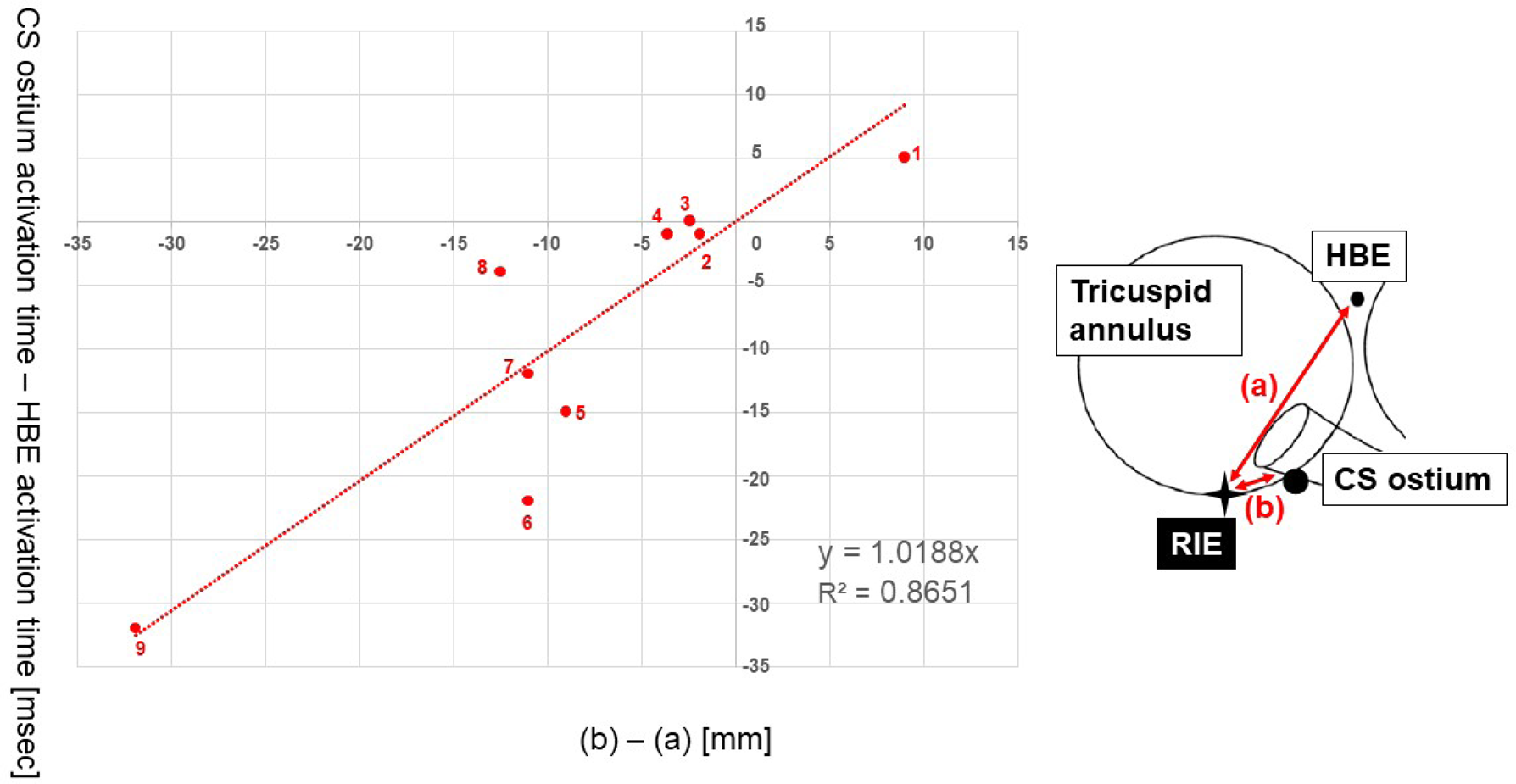
The relationship between activation times and distances of the CS ostium and HBE relative to the RIE reference point. Among 11 AVNRTs with RIE, data on the location was missing in 2. In those 2 AVNRTs, CS ostium was ahead of HBE. In general, the CS ostium was closer to the RIE than the HBE (negative x axis value), and the time to activation after the RIE activation was correspondingly shorter than for the HBE (negative y axis value) in a linear fashion. The graph demonstrates that which of the CS and HBE catheter shows the earlier activation depends on their distances from the atrial exit of RIE. This means that assumption of FP or SP based on which of these two catheters shows earlier activation can be wrong.

### Catheter ablation

In 2 patients who had typical AVNRT as well as atypical AVNRT, conventional SP ablation resulted in inability to induce further AVNRT of any type. The remaining patients received ablation targeting the exit site of LIE and/or RIE. In one of those patients, the tachycardia remained despite ablation of the exit site, and ablation of the conventional SP ablation site was necessary to terminate the tachycardia. In another patient, conventional SP ablation was added after successful exit ablation. Acute success of ablation for AVNRT was achieved in all patients. There were no complications.

The retrograde SP was completely abolished in all patients except for 2 patients, in whom a single AV nodal echo remained. We performed conventional SP ablation in one of them, and in the other patient, a single AV nodal echo through RIE remained even after ablation of both LIE and RIE exits.

During the median follow-up of 2 years, one patient had a recurrence. This patient who received an LIE exit ablation developed a slow/fast AVNRT 2 months later, and had to undergo a second session in which the conventional SP region was ablated.

## Discussion

In this retrospective study of 19 atypical AVNRT in 14 patients, 8 cases had LIE involvement and 11 had RIE. The LIE exits were inside the CS, and localization by 3D mapping and CS electrode catheter matched in 8/8 AVNRTs with LIE. In contrast, RIE exits were on the posterior TA where electrode catheters are conventionally not placed, demonstrating that 3D electroanatomical mapping was necessary for accurate localization. When the RIE exits were involved, the earliest atrial activation site according to conventionally placed electrode catheters was the CS ostium in 7 cases, the His bundle area in 1, and the two sites simultaneously in 3, meaning that the presence of RIE was missed in 4 out of 11 (36%) AVNRTs. Graphing the relationship between activation times and distances demonstrated that which of CS ostium and HBE was activated earlier depended on their distances from the atrial exit of the RIE. This shows that even if the earliest retrograde atrial activation site is not the CS ostium, the retrograde limb of the circuit of atypical AVNRT could still be SP in cases of RIE involvement.

### Retrograde SP

Retrograde dual AV nodal pathways are traditionally diagnosed when VA conduction jumps using the criterion of an increase of at least 50 ms in the VA interval for a 10 ms decrease in the ventricular extrastimulus coupling interval, associated with a change of retrograde atrial activation sequence, namely, a shift in the earliest retrograde atrial activation site from the HBE to the ostium of the CS^2,6^. In the current study, however, we found that the shift from retrograde FP to SP was not necessarily related to the typical retrograde conduction jumps, which was observed in 3 of 6 patients. Moreover, the earliest retrograde atrial activation site among conventionally placed electrode catheters was the CS ostium in 7 among a total of 19 atypical AVNRTs. One of the reasons for these inconsistencies between the conventional way of identifying the SP and our results stems from our ability to identify more anatomically accurate locations of the SP. Conventional wisdom was that the FP lay anteriorly, and SP posteriorly, around the ostium of the CS. Now it is known that the retrograde SP conduction propagates from LIE and/or RIE to the atrium, which are respectively on the mitral annulus and posterior TA^7,8^. Additionally, the extent of change in the retrograde atrial activation sequence from FP to SP conduction may depend on anatomic variation in the size and location of the AV node as well as the magnitude of conduction delay associated with retrograde SP conduction^11^. The position of the HBE catheter electrodes relative to the His bundle may also influence the apparent ordering of activation sequence when distances and activation times are similar.

We believe the criteria for identifying presence of retrograde SP need to be updated based on these latest findings.

### Diagnosing the retrograde pathway of atypical AVNRT

The literature has been indifferent to RIE until recently. The posterior TA area where the RIE lies is a blind spot to electrode catheters, which may be why RIE is ignored. Our current results indicate that which of the CS or HBE catheter activates earlier depends on their distance from the atrial exits of RIE (Figure 5). If this fact isn’t acknowledged, a case such as the example in Figure 4 could easily be misdiagnosed as retrograde FP conduction, although in this study we had only one such patient. Therefore, we recommend placing an electrode catheter covering the posterior TA or employing 3D mapping of that area, to catch possible RIE when atypical AVNRT is suspected (Figure 6).

**Figure 6.**
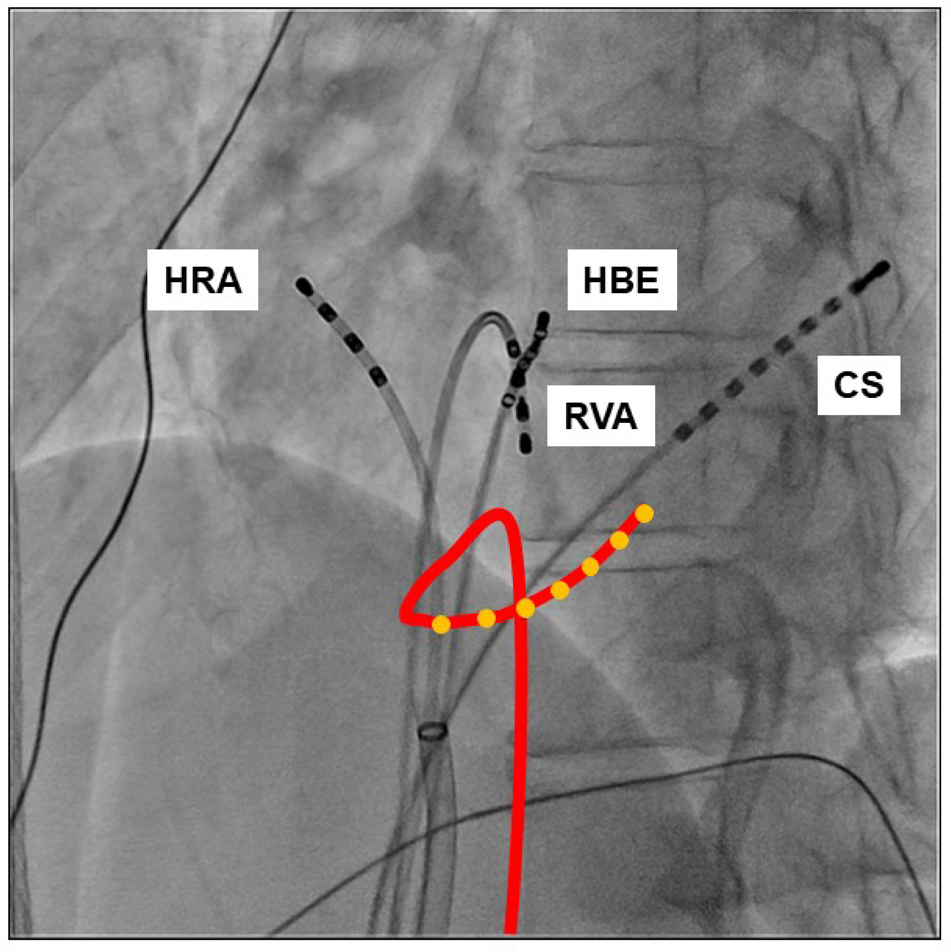
Ideal catheter positioning for RIE exit site identification. To identify the exit site of RIE, we recommend placing a multipolar electrode catheter as shown (red line) in the left anterior oblique (LAO) projection figure, covering the posterior TA from the vicinity of the CS ostium to the isthmus of right atrium. Abbreviations are as in Figure 1.

### Clinical implications

This study revealed that the retrograde SP is different from what has been conventionally defined. It has various exit sites, which can be classified as RIE and LIE. As these diverse exit sites are the earliest atrial sites during retrograde SP conduction, the retrograde SP should be defined as the site having the earliest atrial activation except for the HBE. In fact, in this study there were a few patients in whom the earliest atrial site of retrograde SP was the ostium of CS, as the conventional definition. Instead, there was a variety of retrograde atrial activation patterns.

Our results have led us to change our mapping strategy for atypical AVNRT to intensively target posterior TA with a deliberately placed electrode catheter or electroanatomical 3D mapping (Figure 6).

### Limitations

This study had limitations inherent to any retrospective study. First, it was a single-center trial with a limited number of subjects. EP study could not be done thoroughly in patients in whom AVNRT occurred repeatedly. We were only able to conduct programmed stimulation of the ventricles in 10 out of 14 patients. However, we were able to assess retrograde atrial sequence and earliest site of SP conduction during atypical AVNRT or constant ventricular pacing in all patients. Second, for measurement of distances, we defined CS ostium as its floor. Although the size of the mouth of CS was not measured, it might influence the distances that we have discussed. Third, the inside of the left atrium was not mapped for identifying the exit of LIE. However, catheter ablation targeting the mapped exits in CS succeeded in eliminating retrograde SP conduction via LIE. Lastly, we did not conduct entrainment pacing at the earliest atrial activation sites during tachycardia, therefore, post pacing interval was not examined. The success of ablation at those sites indicated that they were the exits to the atrium of the tachycardia circuits, but strictly speaking, the circuit itself was not proven.

## Conclusion

In atypical AVNRT, conventional electrode catheter placement can expose the existence of LIE with absolute accuracy, but may overlook that of RIE. We believe it essential to place a catheter on or map using a 3D electroanatomical system the posterior TA in cases of suspected atypical AVNRT, so that all inferior extensions of the AV node can be identified and targeted for treatment.

Sources of Funding, & Disclosures

## Data Availability

All data referred to in the manuscript are available.

## References

1. Kawabata M, Maeda S, Kamata T, Kawashima T, Yonai R, Okishige K, Atarashi H, Hirao K. Paradigm Shift for Catheter Ablation of Atypical Atrioventricular Nodal Re-entrant Tachycardia: Three-Dimensional Mapping-Based Ablation. JACC Clin Electrophysiol. 2023;9:1730–1740.

2. Katritsis DG, Marine JE, Latchamsetty R, Zografos T, Tanawuttiwat T, Sheldon SH, Buxton AE, Calkins H, Morady F, Josephson ME. Coexistent types of atrioventricular nodal re-entrant tachycardia: implications for the tachycardia circuit. Circ Arrhythm Electrophysiol. 2015;8:1189–1193.

3. Stavrakis S, Jackman WM, Lockwood D, Nakagawa H, Beckman K, Elkholey K, Wang Z, Po SS. Slow/Fast Atrioventricular Nodal Reentrant Tachycardia Using the Inferolateral Left Atrial Slow Pathway: Role of the Resetting Response to Select the Ablation Target. Circ Arrhythm Electrophysiol. 2018;11:e006631.

4. Nam GB, Rhee KS, Kim J, Choi KJ, Kim YH. Left atrionodal connections in typical and atypical atrioventricular nodal reentrant tachycardias: activation sequence in the coronary sinus and results of radiofrequency catheter ablation. J Cardiovasc Electrophysiol. 2006;17:171–177.

5. Otomo K, Nagata Y, Uno K, Fujiwara H, Iesaka Y. Atypical atrioventricular nodal reentrant tachycardia with eccentric coronary sinus activation: electrophysiological characteristics and essential effects of left-sided ablation inside the coronary sinus. Heart Rhythm. 2007;4:421–432.

6. Gonzalez MD, Branchs JE, Rivera J. Ablation of Atrioventricular Junctional Tachycardias: Atrioventricular Nodal Reentry, Variants, and Focal Junctional Tachycardia. In: Stephen Huang SK, Miller JR, editor. CATHETER ABLATION OF CARDIAC ARRHYTHMIAS. 4th. Philadelphia, USA: Elsevier, 2019:369–408.

7. Inoue S, Becker AE. Posterior extensions of the human compact atrioventricular node: a neglected anatomic feature of potential clinical significance. Circulation. 1998;97:188–193.

8. Anderson RH, Sanchez-Quintana D, Mori S, Cabrera JA, Back Sternick E. Re-evaluation of the structure of the atrioventricular node and its connections with the atrium. Europace. 2020;22:821–830.

9. Knight BP, Ebinger M, Oral H, Kim MH, Sticherling C, Pelosi F, Michaud GF, Strickberger SA, Morady F. Diagnostic value of tachycardia features and pacing maneuvers during paroxysmal supraventricular tachycardia. J Am Coll Cardiol. 2000;36:574–582.

10. Knight BP, Zivin A, Souza J, Flemming M, Pelosi F, Goyal R, Man C, Strickberger SA, Morady F. A technique for the rapid diagnosis of atrial tachycardia in the electrophysiology laboratory. J Am Coll Cardiol. 1999;33: 775–781

11. Hirao K, Otomo K, Wang X, Beckman KJ, McClelland JH, Widman L, Gonzalez MD, Arruda M, Nakagawa H, Lazzara R, et al. Para-Hisian pacing. A new method for differentiating retrograde conduction over an accessory AV pathway from conduction over the AV node. Circulation. 1996;94:1027–1035.

12. Sung RJ, Waxman HL, Saksena S, Juma Z. Sequence of retrograde atrial activation in patients with dual atrioventricular nodal pathways. Circulation. 1981;64:1059–1067.

